# Evaluating the utility of REVEL and CADD for interpreting variants in amyotrophic lateral sclerosis genes

**DOI:** 10.1101/2023.08.03.23293616

**Authors:** Michael R. Fiorini, Allison A. Dilliott, Sali M.K. Farhan

## Abstract

Amyotrophic lateral sclerosis (ALS) is a debilitating neurodegenerative disease affecting approximately two per 100,000 individuals globally. While there are many benefits to offering early genetic testing to people with ALS, this has also led to an increase in the yield of novel variants of uncertain significance in ALS-associated genes. Computational (in silico) predictors, including REVEL and CADD, are widely employed to provide supporting evidence of pathogenicity for variants in conjunction with clinical, molecular, and other genetic evidence. However, in silico predictors are developed to be broadly applied across the human genome; thus, their ability to evaluate the consequences of variation in ALS-associated genes remains unclear. To resolve this ambiguity, we surveyed 20 definitive and moderate ClinGen defined ALS-associated genes from two large, open access ALS sequencing datasets (total people with ALS = 8,230; controls = 9,671) to investigate REVEL and CADD’s ability to predict which variants are most likely to be disease-causing in ALS. While our results indicate a predetermined pathogenicity threshold for REVEL that could be of clinical value for classifying variants in ALS-associated genes, an accurate threshold was not evident for CADD, and both in silico predictors were of limited value for resolving which variants of uncertain significance (VUS) may be likely pathogenic in ALS. Our findings allow us to provide important recommendations of the use of REVEL and CADD scores for variants, and indicate that both tools should be used with caution when attempting to evaluate the pathogenicity of VUSs in ALS genetic testing.

## Introduction

Amyotrophic lateral sclerosis (ALS; [MIM: 105400]) is a neurodegenerative disease characterized by progressive muscle atrophy, weakness, dysarthria, and dysphagia, reflecting adult-onset upper and lower motor neuron degeneration^1^. ALS affects approximately two per 100,000 individuals worldwide, of which 5-10% of cases present with a known family history of disease^1^. Notably, heritability estimates for ALS are high^2^, and mutations known to cause the disease in patients with a family history, including pathogenic variants in *C9orf72* (recurring hexanucleotide repeat expansion; [MIM: 614260]), *SOD1* [MIM: 147450], *TARDBP* [MIM: 605078], and *FUS* [MIM: 13707], have been frequently observed in apparently sporadic patients as well^3–5^. Due to the increasing knowledge of the genetic architecture of ALS, expanding clinical genetic testing beyond those with a family history of disease, as has been the conventional approach, to ensure that all people with ALS carrying clinically actionable variants are identified, has been proposed^6^. Genetic testing is now considered an important tool in the clinical care of people with ALS, as specific genetic profiles may offer early and accurate diagnosis and access to clinical trials^7–9^.

Recent advances of next-generation DNA sequencing (NGS), which encompasses targeted gene panels, exome, and genome sequencing, have allowed rapid and inexpensive genetic sequencing of people with ALS^10^. Yet accurate classification of variant pathogenicity is a significant challenge for all diseases, especially late-onset disorders exhibiting incomplete penetrance. Variant classification typically follows the American College of Medical Genetics and Genomics (ACMG) pathogenicity classification guidelines which relies on multiple lines of distinct evidence, such as functional experimental evidence, minor allele frequency in healthy populations and relevant disease cohorts, and computational (*in silico*) predictors^11^. While some variants can be confidently predicted using these criteria to be either pathogenic or benign, in many cases, clinical and laboratory data can be sparse or even conflicting, and as a result many non-synonymous variants — particularly missense variants — are classified as variants of uncertain significance (VUS). Consequently, VUSs are rapidly accumulating in ALS-associated genes, as shown by genetic testing of known genes in ALS cohorts having identified VUSs in 15-25% of patients^12–15^.

Given the practical limitations of experimentally validating all variants as part of an interpretation workup, computational tools that can accurately predict the pathogenicity of rare variants are often cautiously applied. The incorporation of *in silico* predictors in genetic variant classification was outlined in 2015 by the ACMG, which stated that the lowest level of evidence for pathogenic (PP3-supporting) or benign (BP4-supporting) could be assigned if supported by multiple lines of computational evidence^11^. *In silico* evidence must then be combined with other lines of evidence to classify the variant as being pathogenic, benign, or of uncertain significance.

Many *in silico* predictors have been developed for evaluating PP3/BP4 for missense, splice site, or non-coding variants. Rare Exome Variant Ensemble Learner (REVEL) is an *in silico* method for predicting the pathogenicity of missense variants^16^. REVEL is based on the predictions of 13 tools and provides a score for individual missense variants ranging from 0 to 1, with a higher score reflecting a higher probability that a variant is pathogenic. Combined Annotation-Dependent Depletion (CADD) is an *in silico* method that assigns a score measuring a variant’s deleteriousness, a property that reduces organismal fitness and correlates strongly with molecular functionality and pathogenicity^17–19^. CADD scores are transformed into a Phred-like rank score based on the genome-wide distribution of scores for all ∼9 billion potential single nucleotide variants (3 billion nucleotides and the possible change to the three other types). For instance, a scaled CADD score equal or greater than 10 indicates that a variant is predicted to be among the 10% most deleterious variants, a score equal or greater than 20 indicates that a variant is predicted to be amongst the 1% most deleterious, and so on^18^.

Although REVEL and CADD have both demonstrated remarkable performance on their respective validation datasets, these *in silico* tools are developed to be broadly applied across the human genome and not to specific disease-associated genes. As a result, their ability to accurately predict the consequences of variants in ALS-associated genes remains unclear.

Herein, we investigate REVEL and CADD’s ability to identify pathogenic variants in ALS-associated genes and test the hypothesis that the *in silico* predictors may help resolve VUSs.

## Subjects and Methods

### Participants and genetic sequencing

We obtained sequencing data from the ALS Knowledge Portal^20^ and Project MinE ALS Sequencing Consortium^21^. The ALS Knowledge Portal includes whole exome sequences of 3,864 people with ALS (hereafter referred to as pALS) and 7,839 controls that were subjected to rigorous quality control measures, as previously described, including sequencing depth and coverage assessment at a variant and sample level, genetic ancestry matching by principal component analysis, and relatedness assessment using identity by descent metrics^20^. The Project MinE ALS Sequencing Consortium dataset was obtained from the Project Mine Data Browser and includes whole-genome sequencing data from 4,366 pALS and 1,832 controls. The sequencing methodology and quality control measures closely reflect those applied for the ALS Knowledge Portal and have also been previously described^21^.

### Selection of ALS-associated genes

Sequencing data from the two datasets were restricted to only include variants within 20 known ALS-associated genes (Table S1). Genes were included in the analysis if the gene-disease validity classification from the ALS ClinGen Gene Curation Expert Panel (GCEP)^22^ was definitive or moderate as of July 2023. *C9orf72* was excluded as the pathogenic hexanucleotide repeat expansion was not captured in our analysis.

### Variant annotations and filtering

Variants observed in the datasets were annotated with their respective REVEL^16^ and CADD^19^ scores as well as their ClinVar classification^23^. REVEL (v1.3) scores for all potential missense variants were obtained from ZENODO (DOI 10.5281/zenodo.7072866, accessed February 9, 2023). ClinVar pathogenicity classifications were downloaded from the ClinVar public archive (accessed October 20, 2022) and were binned into six categories: benign/likely benign (B/LB), conflicting significance, VUS, pathogenic/likely pathogenic (P/LP), not observed in ClinVar, and other. Variants binned into the “other” classification included those initially classified as protective, drug response, association, and risk factor/likely risk factor in ClinVar (Table S2). Variants were annotated with CADD scores (v1.6) as well as gene symbols, Genome Aggregation Database (gnomAD; v2.1.1) Non-Finnish European (NFE), non-neurological (n = 51,592) allele frequency and allele counts^24^, and variant consequence using the Variant Effect Predictor (VEP; v.109.0)^25^. The transcript identifiers used as input into VEP for each dataset are provided in Table S3. Coding variant consequences were re-classified into three categories: 1) protein-truncating variant (PTV), 2) missense, and 3) synonymous. PTVs included those classified as: “splice_donor_variant”, “splice_acceptor_variant”, “frameshift_variant”, and “stop_gained”.

Finally, to focus our analyses on rare, coding variants in ALS-associated genes, we filtered the ALS Knowledge Portal and Project MinE ALS Sequencing Consortium datasets to include PTV and missense variants that were rare in the general population (allele frequency < 0.01 in the gnomAD v2.1.1 NFE non-neurological cohort). Because REVEL can only be applied to evaluate missense variants, we focused our score-specific analyses for both tools on missense variants only.

### Assessing REVEL and CADD scores according to ClinVar classifications

To determine whether REVEL and CADD scores were lower, on average, for rare, missense variants in ALS-associated genes classified as B/LB in ClinVar than other pathogenicity classifications, we conducted Wilcoxon Rank-sum tests comparing the mean scores of each ClinVar category – Conflicting, VUS, and P/LP – to the mean score of variants classified as B/LB in ClinVar.

To assess the agreement between REVEL and CADD scores across rare, missense variants in ALS-associated genes, we measured the correlation between the scores across all unique missense variants seen in both datasets using Pearson’s method. We repeated this analysis following the stratification of unique missense variants by ClinVar classification.

### Assessing REVEL and CADD pathogenicity thresholds for variants in ALS-associated genes

We used ClinVar classifications to define variant pathogenicity and established binary variant categories by only including missense variants classified as P/LP and B/LB in ClinVar; P/LP variants were set as the positive reference, while B/LB variants were set as the negative reference. We then fit binary logistic regression models to compute the odds that a variant was classified as P/LP in ClinVar given a particular REVEL or CADD score, as shown in equations 1 and 2 for REVEL and CADD, respectively.

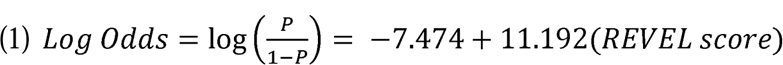

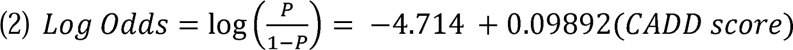

To assess the performance of REVEL and CADD for predicting missense P/LP variants, we harnessed the binary logistic regression models to compute receiver operating characteristic (ROC) curves using the empirical method implemented in the *ROCit* (v2.1.1) R package^26^. To compare the usefulness of REVEL and CADD for predicting missense P/LP variants, we computed the area under the empirically estimated ROC curves (AUC); a higher AUC suggests that the tool performs better at distinguishing between the positive – P/LP variants in ClinVar – and negative – B/LB variants in ClinVar – classes. Then, to identify the score thresholds for which the maximum difference was seen between the true positive rate (TPR; sensitivity) and false positive rate (FPR; 1 – specificity), we computed the maximum Youden index (J) for the empirically estimated ROC curves. In addition, we calculated the accuracy, sensitivity, specificity, positive predictive value (PPV), and negative predictive value (NPV) of REVEL and CADD again using the *ROCit* (v2.1.1) R package.

We produced Kolmogorov-Smirnov plots using the *ROCit* (v2.1.1) R package^26^. Specifically, the cumulative empirical distribution functions for missense B/LB and P/LP variants were plotted across REVEL and CADD scores. The value of the cumulative empirical distribution at any specific REVEL or CADD score was the fraction of variants – B/LB or P/LP in ClinVar – with scores less than or equal to the specified REVEL or CADD score. We then identified the REVEL and CADD score at which the greatest distance was seen between the empirical cumulative distributions of B/LB and P/LP variants and computed the Kolmogorov-Smirnov statistic as the cumulative empirical distribution of B/LB variants subtracted from the cumulative empirical distribution of P/LP variants.

### Quantifying the enrichment of VUS in pALS and controls

To determine whether pALS were enriched for missense VUS that exceeded REVEL or CADD’s respective pathogenicity thresholds compared to controls, we applied a two-sided Fisher’s exact test. Rare, missense VUS in ALS-associated genes were binned based on REVEL’s^27^ and CADD’s^18^ predefined pathogenicity thresholds (REVEL: 0.000-0.643, ≥ 0.644; CADD: 0.000-19.999, ≥20.000) and the proportion of observed variants in pALS from each bin were compared to the proportion of observed variants in controls from each bin, for the ALS Knowledge Portal and Project MinE ALS Sequencing Consortium datasets independently. We also applied a combined analysis across the two cohorts using the Cochran-Mantel-Haenszel (CMH) test. For these analyses, VUS included variants characterized as either VUS in ClinVar, conflicting in ClinVar, or absent from ClinVar.

### Variant-level assessments of REVEL and CADD scores

We compared REVEL and CADD scores of missense variants observed in pALS and controls at a variant level. To determine whether REVEL and CADD scores were correlated with variant odds ratio we computed Pearson’s correlation. We calculated variant odds ratios for missense variants seen across both datasets using equation 3:

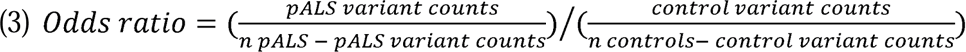

Findings of the odds ratio analysis were validated using Pearson’s correlation and pALS carrier ratio. The pALS carrier ratio for missense variants observed in pALS across both datasets was calculated using equation 4, as well as the gnomAD v2.1.1 NFE, non-neurological allele counts^24^, which offered a control cohort proxy of larger sample size than the ALS Knowledge Portal and Project MinE ALS sequencing consortium.

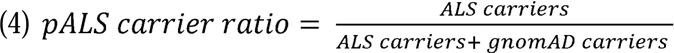

To determine whether using REVEL or CADD could uniquely identify missense VUS of potential interest in pALS but not in controls, we applied a variant-level yield analysis and compared the proportion of observed variants in pALS and controls identified as supporting pathogenic. Rare, coding variants in ALS-associated genes seen in pALS and controls across both datasets were considered supporting pathogenic if they were: P/LP in ClinVar, VUS in ClinVar and PTV, VUS in ClinVar and missense with REVEL ≥ 0.644/CADD ≥ 20.000, absent from ClinVar and PTV, or absent from ClinVar and missense with REVEL ≥0.644/CADD ≥20.000.

### Statistical analyses

We performed all statistical analyses using the R statistical software (v4.1.1)^28^ in R studio (v1.4.1717). In addition, we used the *ggplot2* R package (v3.4)^29^ for data visualization.

## Results

### Distribution of variants in ALS-associated genes

A summary of the ALS Knowledge Portal (3,864 pALS and 7,839 controls) and Project MinE (4,366 pALS and 1,832 controls) datasets used in our study is shown in Figure 1A. For these datasets, we identified 1,395 and 1,149 variants from 20 ClinGen definitive and moderate ALS genes. No variants were observed in *CHCHD10* [MIM: 615903] in the ALS Knowledge Portal (poorly covered in exomes). Similarly, no variants were observed in *UBQLN2* [MIM: 300264] in Project MinE (X-linked gene, not provided). As expected, the proportion of observed variants (synonymous, missense, and PTV) were similar between both datasets for pALS and controls, with a greater frequency of PTVs in cases (Figure 1B). Further, of the 2,180 unique variants from both datasets, 49.69% were previously reported in the ClinVar database (Figure 1C). Among the 9,833 observed variants across both datasets in pALS and controls, a marginally greater proportion of B/LB variants was observed in controls (43.91% and 48.25% of variants in pALS and controls, respectively), while a greater proportion of P/LP variants was observed in pALS (4.74% and 0.846% of variants in pALS and controls, respectively) (Figure 1D). Notably, 46.91% and 47.22% of observed variants in pALS and controls, respectively, were classified as VUS, conflicting, or were absent from ClinVar.

**Figure 1.**
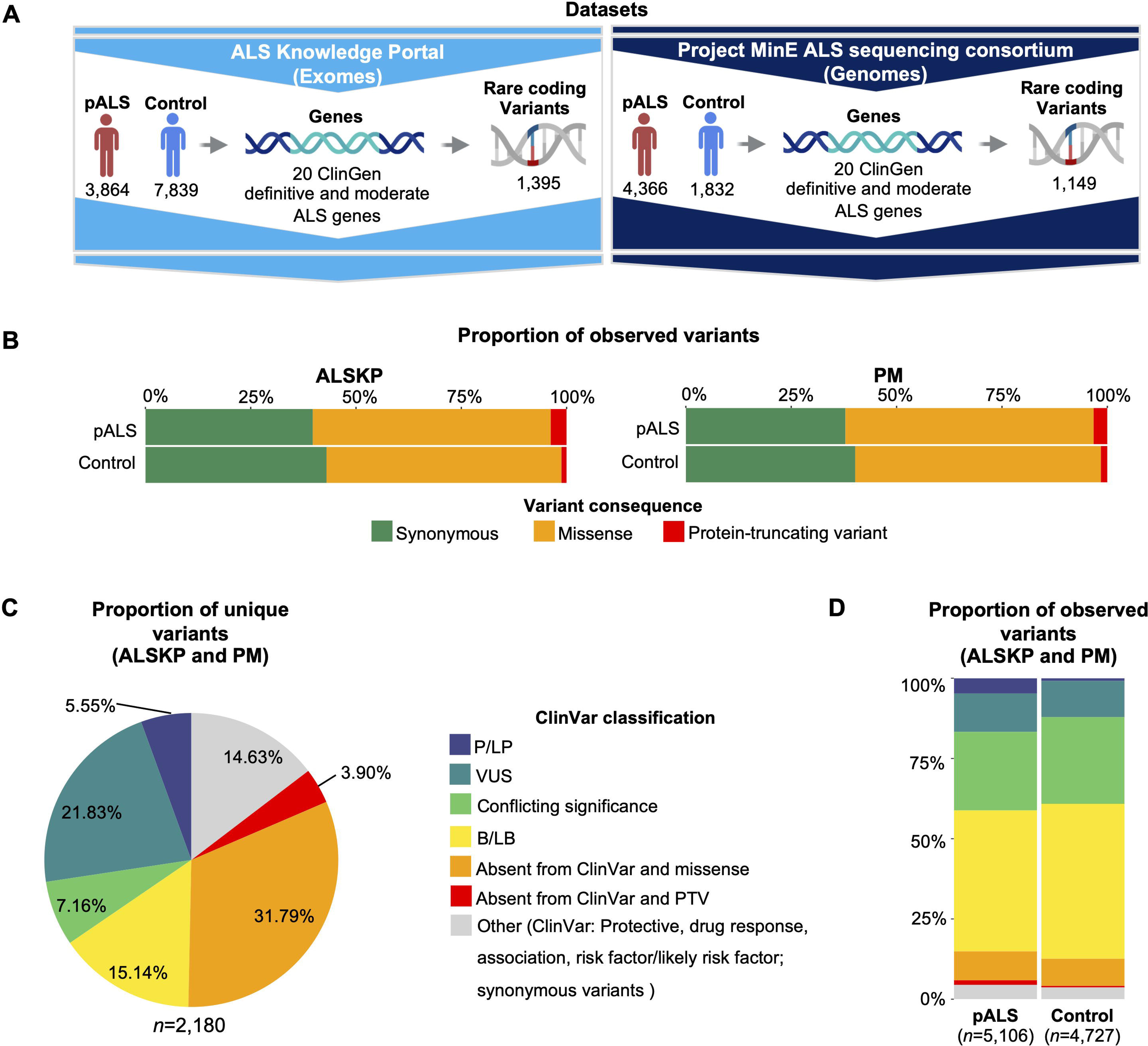
Summary of the ALSKP and PM datasets. A) Description of the sample sizes and rare, coding variants identified in ALS-associated genes in the ALS Knowledge Portal (ALSKP) and Project Mine ALS Sequencing Consortium (PM) datasets. No variants were observed in *CHCHD10* (poorly covered in exomes) in the ALSKP dataset, while no variants were observed in *UBQLN2* (X-linked gene, not provided) in PM dataset. **B)** Proportion of observed variants in the ALSKP and PM datasets according to variant consequence, stratified by ALS status. **C)** Proportion of unique variants observed across the ALSKP and PM datasets categorized by ClinVar classification. **D)** Proportion of observed variants across the ALSKP and PM datasets categorized by ClinVar classification and stratified by ALS status. The variant category denoted as “other” includes variants classified as protective, drug response, association, risk factor/likely risk factor in ClinVar, and all synonymous variants. Abbreviations: B/LB, benign/likely benign; pALS, people with ALS; P/LP, pathogenic/likely pathogenic; PTV, protein-truncating variant; VUS, variant of uncertain significance.

### Accuracy of REVEL and CADD for missense variants in ALS-associated genes

To evaluate the accuracy of REVEL and CADD pathogenicity estimation for missense variants in ALS-associated genes, we investigated the relationship between the scores and ClinVar classifications. The mean REVEL score for B/LB variants (0.237) differed significantly from P/LP variants (0.713; P = 5.43e-15) and VUS (0.319; P = 2.56e-2), but not from variants of conflicting significance (0.304; P = 1.87e-1) (Figure 2A; Figure S1). Similarly, the mean CADD score for B/LB variants (18.650) differed significantly from variants classified as P/LP (24.461; P = 2.61e-6) and VUS (20.966; P = 3.59e-2), but not from variants of conflicting significance (19.032; P = 7.56e-1) (Figure 2B; Figure S1). Similar results were observed in the distribution of REVEL and CADD scores per ClinVar classifications on a per gene-basis (Figure S2).

**Figure 2.**
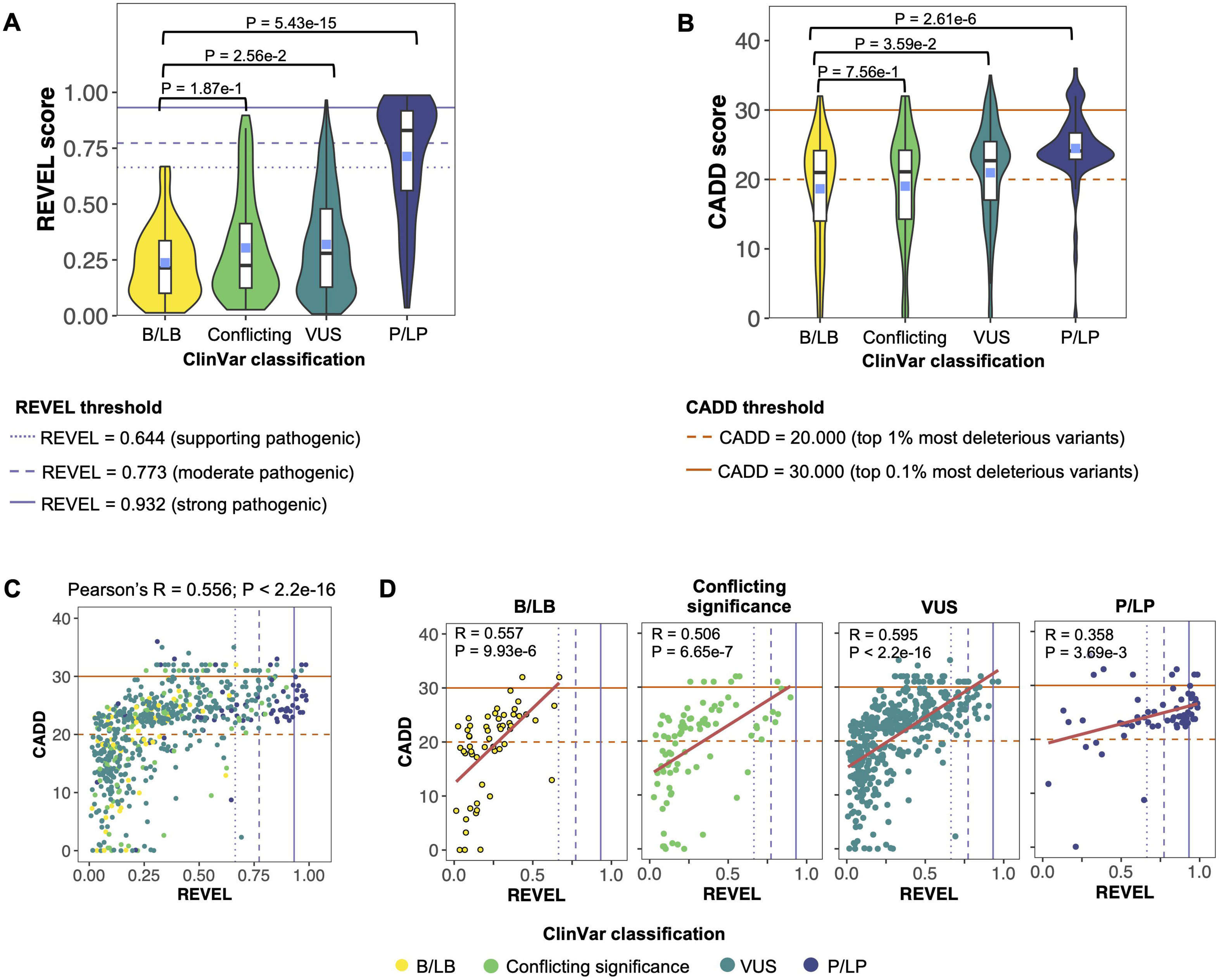
REVEL and CADD scores of rare, missense variants in ALS-associated genes categorized by ClinVar classification. A) REVEL scores of the unique missense variants identified in ALS-associated genes in the ALS Knowledge Portal (ALSKP) and Project Mine ALS Sequencing Consortium (PM) datasets, stratified by ClinVar classification. **B)** CADD scores of the unique missense variants identified in ALS-associated genes in the ALSKP and PM datasets, stratified by ClinVar classification. Wilcoxon Rank-sum tests were conducted to compare the mean REVEL or CADD scores of each ClinVar category to the mean REVEL or CADD score of benign/likely benign (B/LB) variants. **C)** Pearson’s correlation between CADD and REVEL scores of unique missense variants identified in the ALSKP and PM datasets. **D)** Pearson’s correlation between CADD and REVEL scores of unique missense variants identified in the ALSKP and PM datasets, stratified by ClinVar classification. Abbreviations: P/LP, pathogenic/likely pathogenic; VUS, variant of uncertain significance.

Based on the tools’ predetermined pathogenicity cut-offs, we categorized REVEL scores into three bins: 0.000-0.182, 0.183-0.643, and ≥0.644, and CADD scores into three bins: 0.000-9.999, 10.00-19.999, and ≥20.000 to assess the ClinVar classifications of the binned variants (Figure S3)^18, 27^. One missense B/LB variant (1.69%) had a REVEL score ≥ 0.644, while 31 of 59 missense B/LB variants (52.54%) had CADD scores ≥ 20.000. Further, 43 of 65 P/LP missense variants (66.15%) had REVEL scores ≥ 0.644, while 59 of 65 missense P/LP variants (90.77%) had CADD scores ≥ 20.000. Regardless of ClinVar classification and including those absent from ClinVar, 68.24% of unique missense variants had CADD scores ≥ 20.000.

To evaluate the concordance between REVEL and CADD scores in ALS-associated genes, we measured the correlation between the two scores across all unique missense variants observed in either sequencing dataset and found they indeed demonstrated a statistically significant, positive correlation (Pearson’s R = 0.556, P-value < 2.2e-16; Figure 2C). We repeated this exercise following stratification of variants by ClinVar classification, and consistently observed statistically significant positive correlations (Figure 2D).

Additionally, we determined the REVEL and CADD thresholds at which pathogenicity may be most accurately defined for variants in ALS-associated genes. Here, we used ClinVar classifications to define variant pathogenicity and only included missense variants that were classified as B/LB and P/LP. A Kolmogorov-Smirnov test was used to identify the CADD and REVEL scores that described the maximum distance (D) between the empirical cumulative distributions of missense variants classified as B/LB and P/LP in ClinVar (Figure 3A). A REVEL score of 0.458 showed the maximum distance between B/LB and P/LP variants (D = 0.806, P-value < 2.2e-16), while a CADD score of 22.42 showed the maximum distance between B/LB and P/LP variants (D = 0.543, P-value < 2.2e-16). Next, an ROC curve was used to examine the relationship between the true positive rate (TPR) and false positive rate (FPR) for each tool; REVEL showed a high AUC of 0.929 while CADD showed a moderate AUC of 0.754. REVEL had a Youden index at 0.493, indicating the scores resulting in the maximum difference between the FPR and TPR and corresponding to an FPR of 0.032 and a TPR of 0.838 (Figure 3B). CADD had a Youden index at 22.501, corresponding to an FPR of 0.310 and a TPR of 0.853. We then plotted the positive predictive value (PPV) (Figure 3C), negative predictive value (NPV) (Figure 3D), accuracy (Figure 3E), specificity (Figure S4A), and sensitivity (Figure S4B) for REVEL and CADD. REVEL had high PPVs at pathogenicity thresholds of 0.644 (PPV = 0.970), 0.773 (PPV = 1.00), and 0.932 (PPV = 1.00). In contrast, CADD had low PPVs at pathogenicity thresholds of 20.000 (PPV = 0.101) and 30.000 (PPV = 0.052). REVEL also had high NPVs at pathogenicity thresholds of 0.644 (NPV = 0.977), 0.773 (NPV = 0.967), and 0.932 (NPV = 0.940). Similarly, CADD had high NPVs at pathogenicity threshold of 20.000 (NPV = 0.983) and 30.000 (NPV = 0.923).

**Figure 3.**
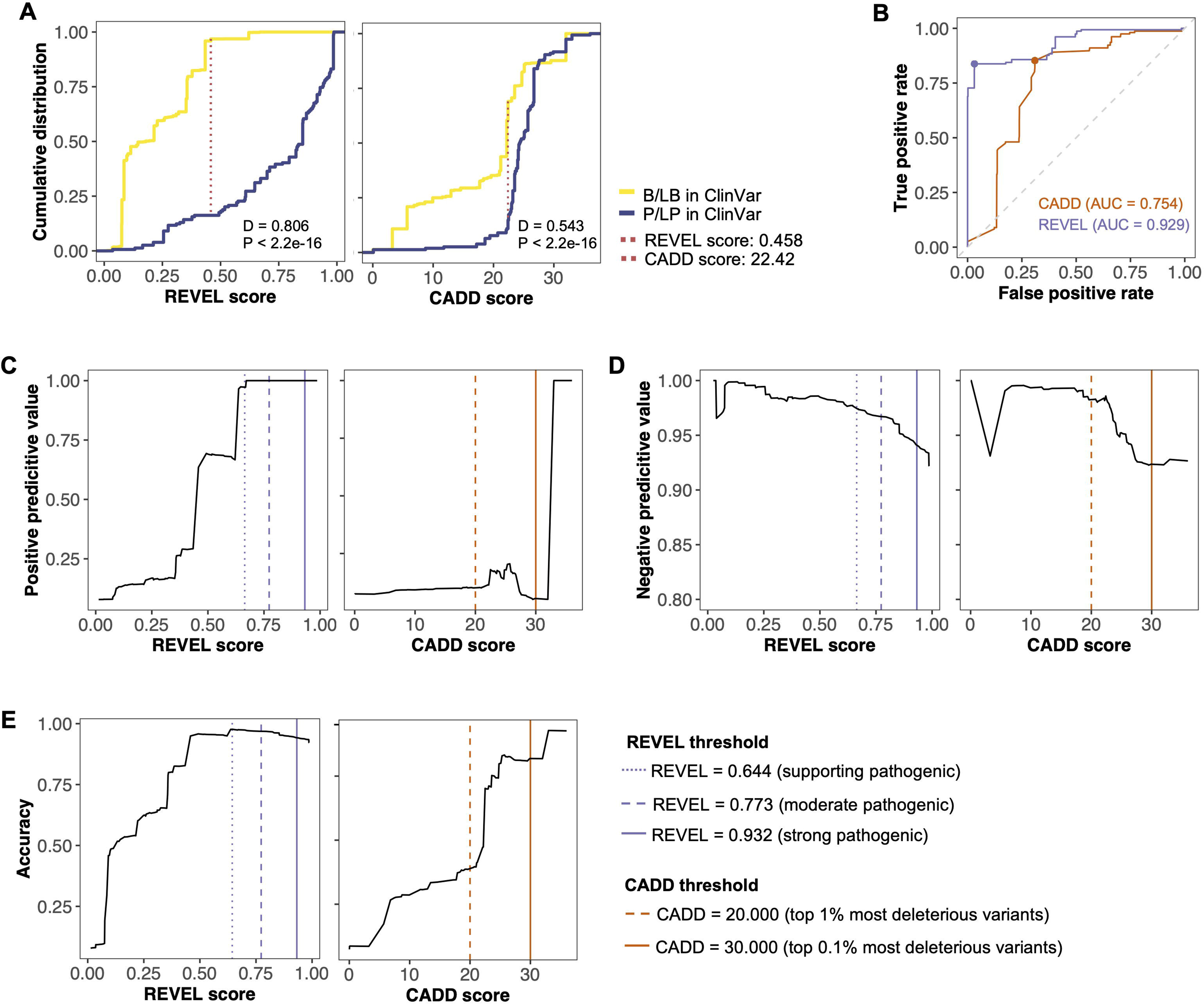
Evaluation of REVEL and CADD scores for rare, missense variants in ALS- associated genes. Performance evaluations were conducted on rare, missense variants in ALS- associated genes classified as benign/likely benign (B/LB) or pathogenic/likely pathogenic (P/LP) in ClinVar **A)** Kolmogorov-Smirnov plots for REVEL and CADD. The red, vertical line represents the REVEL or CADD score that describes the maximum distance between the empirical cumulative distribution of missense variants classified as B/LB and P/LP in ClinVar. **B)** Receiver operating characteristic (ROC) curve and area under the curve (AUC) for REVEL and CADD. The purple and orange points represent the Youden index for REVEL and CADD, respectively. **C)** Positive predictive value of REVEL and CADD. **D)** negative predictive value of REVEL and CADD. **D)** Accuracy of REVEL and CADD.

These results suggest that pathogenicity for rare, coding variants in ALS-associated genes is best defined at a pre-determined threshold of 0.644 for REVEL and 20.000 for CADD. Subsequent analyses employed these pathogenicity thresholds to evaluate REVEL and CADD’s ability to assess the potential pathogenicity of variants in ALS-associated genes characterized as either VUS in ClinVar, conflicting in ClinVar, or absent from ClinVar, hereafter collectively referred to as VUS.

### Leveraging REVEL and CADD to resolve the pathogenicity of missense VUS in ALS-associated genes

We next aimed to determine REVEL and CADD’s ability to evaluate the pathogenicity of missense VUS in ALS-associated genes. Across both datasets, pALS had higher proportions of observed missense VUS than controls with a REVEL score ≥ 0.644 (pALS = 9.69%, controls = 7.26%; Figure 4A). In contrast, across both datasets, controls had a marginally higher proportion of observed missense VUS with a CADD score of 20.000-30.000 than pALS (pALS = 49.59%, controls = 52.15%), but pALS had a higher proportion of missense VUS with a CADD score ≥30.000 than controls (pALS = 13.00%, controls = 11.27%; Figure 4A). Similar results were observed when both datasets were analyzed independently (Figures S5A and S5B).

**Figure 4.**
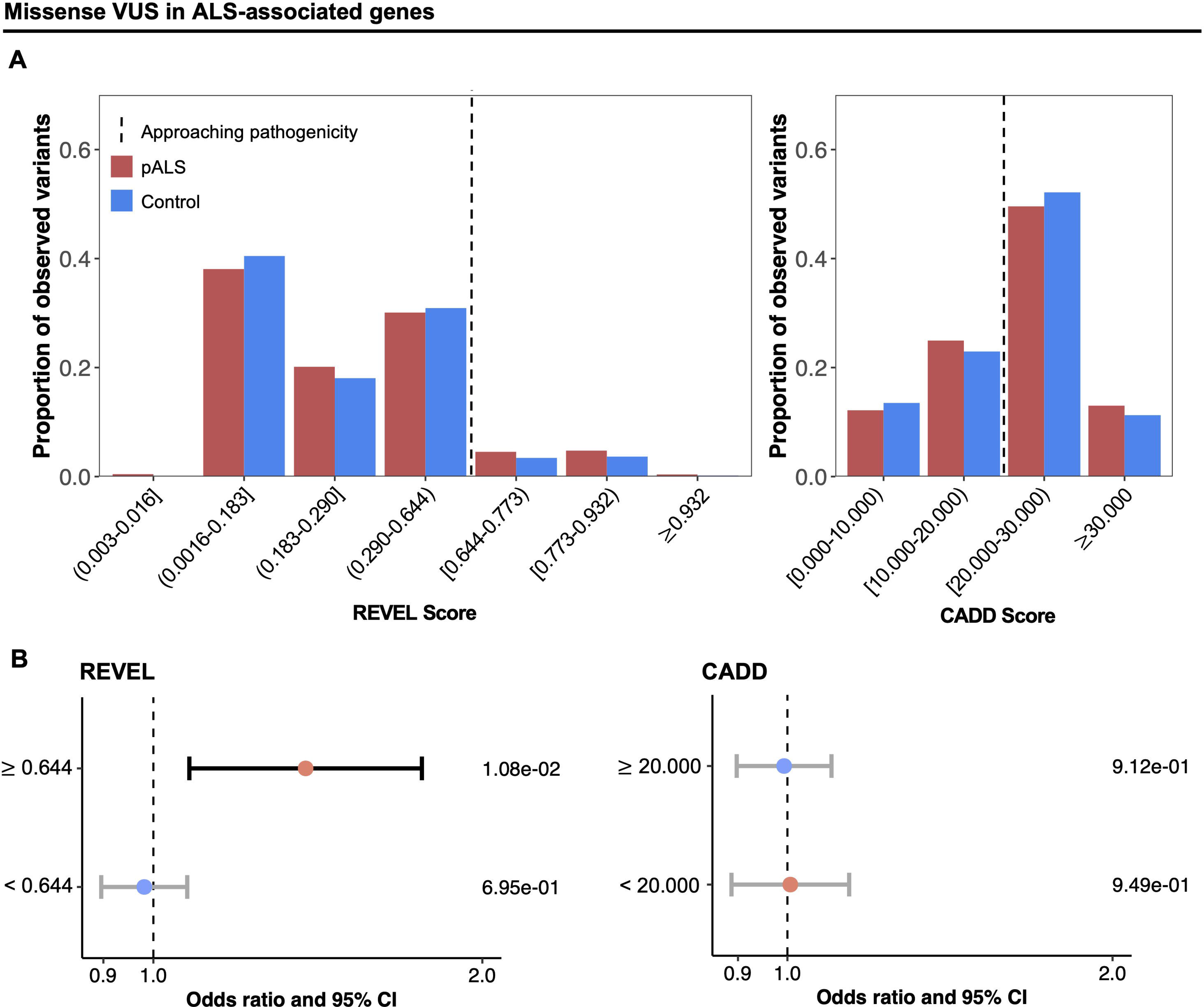
Distribution of rare, missense VUS in ALS-associated genes. Missense variants categorized as variants of uncertain significance (VUS) in ClinVar, conflicting in ClinVar, or absent from ClinVar are collectively referred to as VUS. **A)** Proportion of observed VUS across the ALS Knowledge Portal and Project MinE ALS Sequencing Consortium (PM) datasets, stratified by ALS status and binned REVEL or CADD score. **B)** Cochran-Mantel-Haenszel (CMH) tests investigating whether people with ALS (pALS) were enriched for VUS that had REVEL or CADD scores that exceeded their respective pathogenicity thresholds (REVEL ≥ 0.644; CADD ≥ 20.000) compared to controls. P-values corresponding to each variant category are shown on the right of the plot. Abbreviations: CI, confidence interval.

We investigated whether pALS were enriched for VUS that had REVEL or CADD scores that exceeded their respective pathogenicity thresholds. The analyses revealed that pALS were indeed enriched for VUS with a REVEL score ≥ 0.644 (OR = 1.378 [1.079-1.761], P = 1.08e-2), but not for VUS with a REVEL score < 0.644 (OR = 0.981 [0.896-1.074], P = 6.95e-1), across the combined dataset (Figure 4B). In contrast, pALS were not enriched for VUS that had a CADD score ≥ 20.000 (OR = 0.993 [0.898-1.098], P = 9.12e-1), nor VUS that had a CADD score < 20.000 (OR = 1.006 [0.888-1.141], P = 9.49e-1), across the combined dataset (Figure 4B). These results again reflected those observed when both datasets were analyzed independently (Figures S5C and S5D).

Next, we examined the correlation between the *in silico* scores and variant odds ratio for rare, missense variants in ALS-associated genes. Across all variants, REVEL and CADD scores both demonstrated a positive correlation with variant odds ratio (R = 0.23, P < 2.2e-16 and R = 0.058, P = 1.8e-5, respectively; Figures 5A and 5B). Similarly, when we restricted the analyses to only include variants classified as VUS or conflicting significance in ClinVar, REVEL and CADD both maintained weak positive correlations with variant odds ratio (R = 0.053, P = 8.8e-3 and R = 0.053, P = 8.8e-3, respectively; Figures 5C and 5D). We also examined whether the scores were correlated with pALS carrier ratio – respective to gnomAD v2.1.1 NFE, non-neurological allele counts^24^ – to validate the findings of the odds ratio analyses. Again, across all variants both REVEL and CADD demonstrated statistically significant positive correlations between their respective scores and pALS carrier ratio (R = 0.31, P < 2.2e-16 and R = 0.094, P = 2.4e-12, respectively; Figures 5E and 5F), which was maintained when restricted to variants classified as VUS or conflicting significance in ClinVar (R = 0.14, P = 1.3e-12 and R = 0.07, P = 5.5e-4, respectively; Figures 5G and 5H). The correlations between REVEL or CADD score and variant odds ratio and pALS carrier ratio for missense variants classified as P/LP and B/LB in ClinVar, as well as variants absent from ClinVar are shown in Figures S5 and S6.

**Figure 5.**
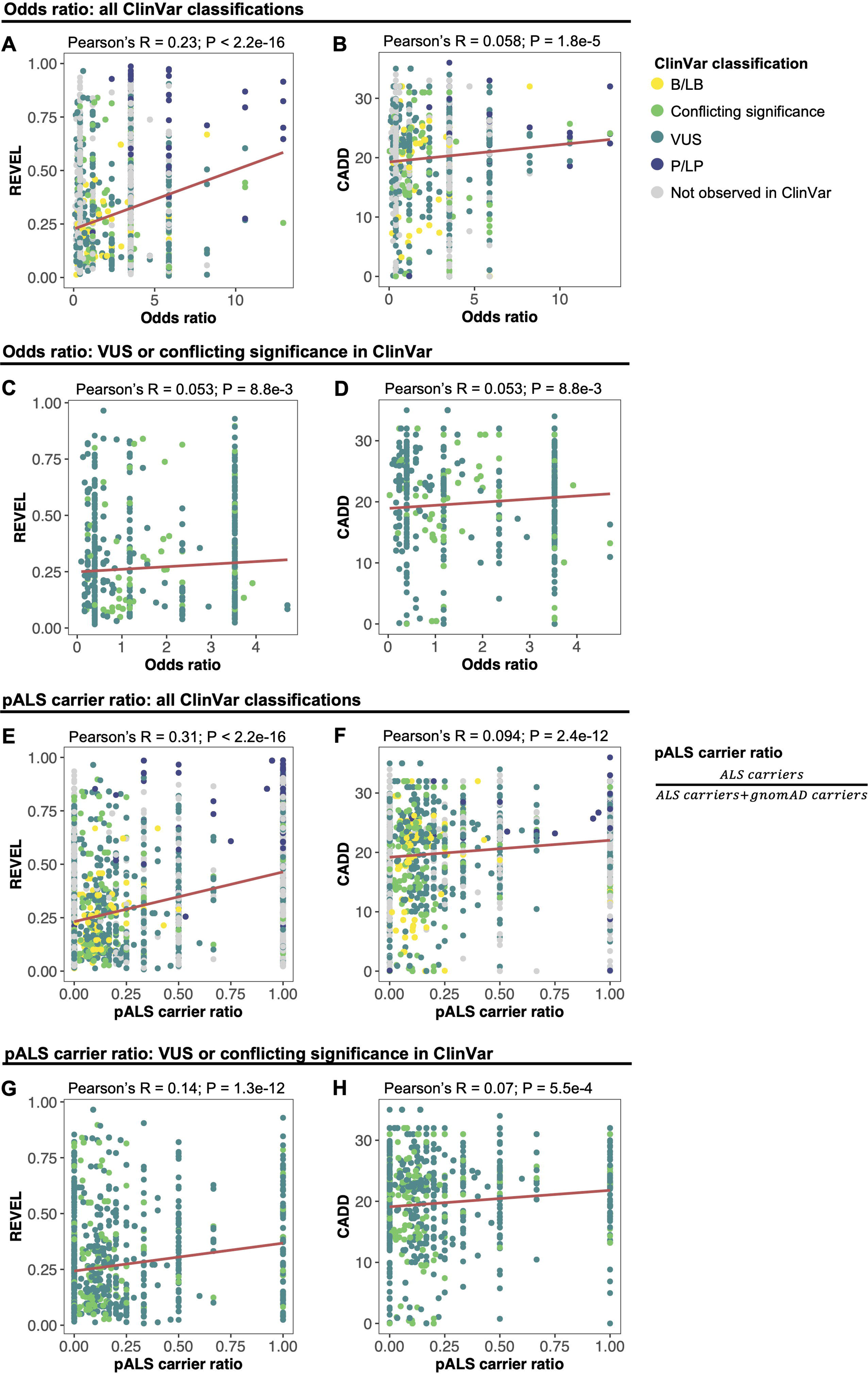
REVEL and CADD scores according to variant odds ratio and pALS carrier ratio. Pearson’s correlation was computed using rare, missense variants in ALS-associated genes. **A)** Correlation between REVEL score and variant odds ratio across all ClinVar categories. **B)** Correlation between CADD score and variant odds ratio across all ClinVar categories. **C)** Correlation between REVEL score and variant odds ratio for variants of uncertain significance (VUS) and conflicting significance in ClinVar. **D)** Correlation between CADD score and variant odds ratio for VUS and conflicting significance in ClinVar. **E)** Correlation between REVEL score and variant carrier ratio in people with ALS (pALS carrier ratio) across all ClinVar categories. **F)** Correlation between CADD score and pALS carrier ratio across all ClinVar categories. **G)** Correlation between REVEL score and pALS carrier ratio for VUS and conflicting significance in ClinVar. **H)** Correlation between CADD score and pALS carrier ratio for VUS and conflicting significance in ClinVar. Only variants that had an odds ratio within three standard deviations of the mean are shown and were included when computing Pearson’s correlation. Abbreviations: B/LB, benign/likely benign; P/LP, pathogenic/likely pathogenic.

**Figure 6.**
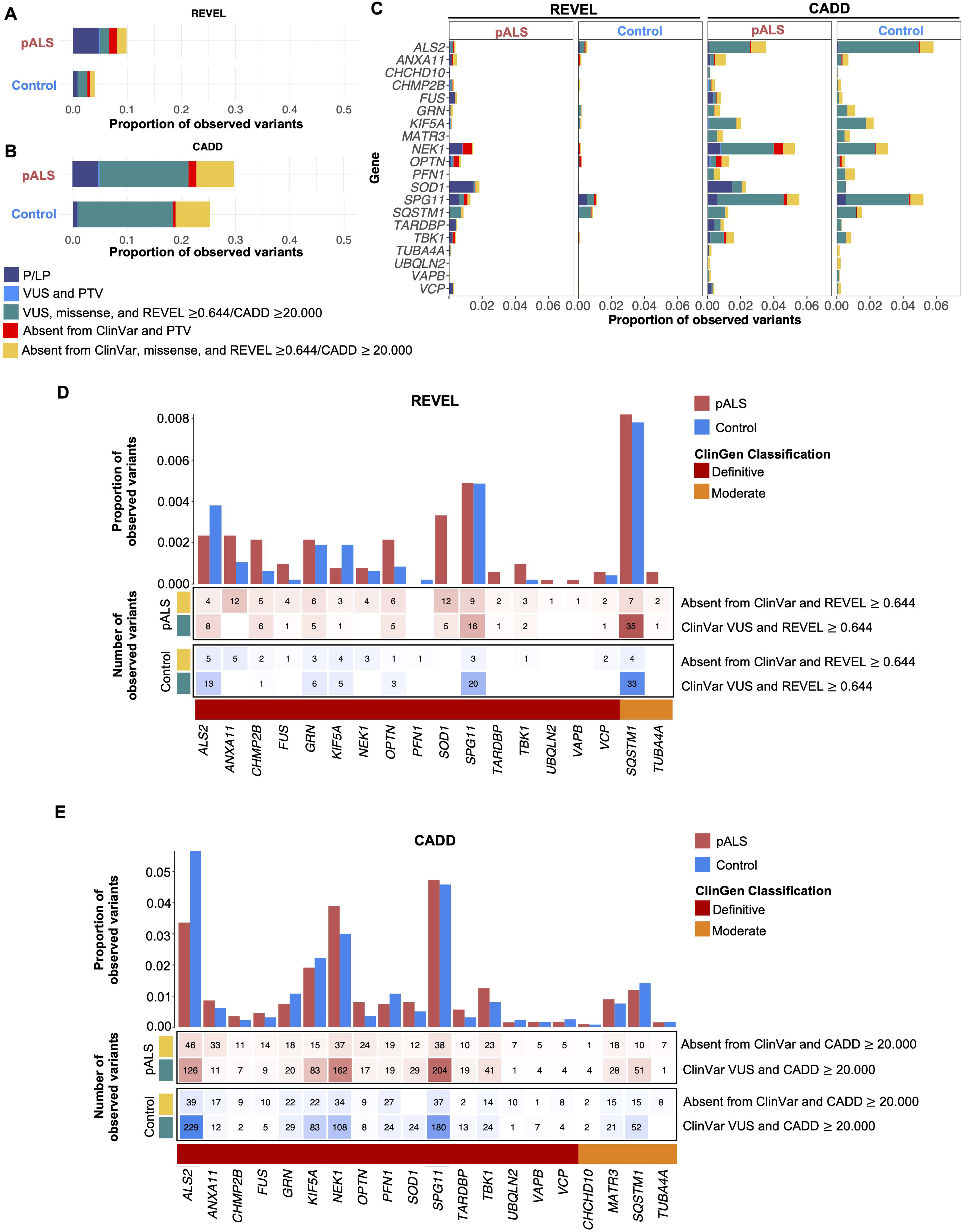
Harnessing REVEL and CADD to identify supporting pathogenic rare, missense variants in ALS-associated genes. Variants were considered supporting pathogenic if they were either pathogenic/likely pathogenic (P/LP) in ClinVar, protein-truncating variant (PTV) of uncertain significance in ClinVar, missense variant of uncertain significance (VUS) in ClinVar with REVEL ≥ 0.644/CADD ≥ 20.000, PTV absent from ClinVar, or a missense variant absent from ClinVar with REVEL ≥ 0.644/CADD ≥ 20.000. The proportion of observed missense variants in pALS and controls from the ALS Knowledge Portal (ALSKP) and Project MinE ALS Sequencing Consortium (PM) datasets that were identified as supporting pathogenic by **A)** ClinVar and REVEL and **B)** ClinVar and CADD are presented. **C)** Supporting pathogenic missense variants seen in pALS and controls across the ALSKP and PM datasets, stratified by ALS status and ALS-associated gene. **D)** Missense variants identified in the ALSKP and PM datasets that were either VUS in ClinVar with REVEL ≥ 0.644 or absent from ClinVar with REVEL ≥ 0.644, stratified by ALS status and ALS-associated gene. **E)** Missense variants identified in the ALSKP and PM datasets that were either VUS in ClinVar with CADD ≥ 20.000 or absent from ClinVar with CADD ≥ 20.000, stratified by ALS status and ALS-associated gene. The heatmaps show the number of observed variants seen in people with ALS (pALS) and controls while the bar charts show the proportion of observed variants seen in pALS and controls. The colored track beneath the heatmap represents the gene-disease validity classification from the ALS ClinGen Gene Curation Expert Panel^22^ for the corresponding gene.

We applied a variant-level yield analysis to determine whether using REVEL or CADD may identify missense VUS of potential interest unique to pALS. Of the 5,106 rare, coding variants observed in pALS from the combined dataset, 328 variants (6.42%) were P/LP in ClinVar, VUS in ClinVar but are protein-truncating variant (PTV), or absent from ClinVar and PTV (Figures 6A and 6B; Table S4). In contrast, 66 of these types of variants (1.40%) were observed in controls. In addition, REVEL scores indicated that 175 missense variants (3.43%) in pALS and 123 missense variants (2.60%) in controls were supporting pathogenic, including missense VUS in ClinVar with REVEL ≥ 0.644 and missense variants absent from ClinVar with REVEL ≥ 0.644. Finally, 1,193 variants (23.36%) in pALS and 1,129 variants (23.88%) in controls had CADD scores ≥ 20.000, including missense VUS in ClinVar and missense variants absent from ClinVar. The independent variant-level yield analyses for each dataset are shown in Figure S7 and reflect the results observed in the merged analysis.

A higher proportion of REVEL-defined supporting pathogenic missense VUS were observed in pALS than controls for 15 of 18 genes observed in both datasets. The greatest number of REVEL- or ClinVar-defined supporting pathogenic missense variants in pALS were observed in *NEK1* [MIM: 604588], *SOD1*, and *SPG11* [MIM: 610844], while *ALS2* [MIM: 606352],

*SQSTM1* [MIM: 601530], and *SPG11* harbored the greatest number of supporting pathogenic missense variants in controls (Figure 6C). Notably, both *ALS2* and *SPG11* are large multi-exon recessive genes. There were also REVEL-defined supporting pathogenic missense variants observed exclusively in pALS in *SOD1* (17 observed variants), *TUBA4A* (3 observed variants; [MIM: 191110]), *UBQLN2* (1 observed variant), and *VAPB* (1 observed variant; [MIM: 605704]) (Figure 5D). Analyzing the ≥

20.000 CADD missense VUS revealed that 12 of 20 genes showed a higher proportion of observed variants in pALS than controls (Figure 6E), yet *ALS2, NEK1,* and *SPG11* harbored the greatest number of CADD- or ClinVar-defined supporting pathogenic missense variants in both pALS and controls (Figure 6C). There was also a selection of genes with surprisingly high frequencies of CADD ≥20.000 within controls, including *ALS2* (268 observed variants), *KIF5A* (105 observed variants; [MIM: 602821]), *NEK1* (142 observed variants), *SPG11* (217 observed variants). These findings should be interpreted in the context of gene size, inheritance pattern, and incomplete penetrance.

## Discussion

The increasing number of novel variants reported in ALS-associated genes has created new challenges in standardized clinical testing in ALS. Computational (*in silico)* predictors, including REVEL and CADD, are widely employed in clinical and research settings to provide supporting evidence of pathogenicity^30^. However, *in silico* predictors are developed to be broadly applied across the human genome and not to specific diseases or genes^27^. As a result, their ability to evaluate the consequences of rare, missense variants in ALS-associated genes remains unclear and could lead to misinterpretation. To resolve this uncertainty, we extracted 20 genes classified as ‘definitive’ or ‘moderate’ ALS genes as evaluated by the ALS ClinGen GCEP, from two pALS-control open access, sequencing datasets to investigate REVEL and CADD’s ability to predict which missense variants are contributing to disease pathogenesis. While our results indicate a predetermined pathogenicity cut-off for REVEL that could be of clinical value for classifying variants in ALS-associated genes, an accurate cut-off was not evident for CADD, and both *in silico* predictors were of limited value for resolving which VUS are truly pathogenic in ALS. Our findings allow us to provide important recommendations for interpreting REVEL and CADD scores for missense variants in ALS-associated genes and indicate that both tools should be used with caution when attempting to evaluate the pathogenicity of VUS in ALS genetic testing.

A major limitation of *in silico* predictors is that score thresholds are typically established using a genome-wide set of variants and, as a result, are often not calibrated to particular genes or diseases^27^. In our analysis, we leveraged REVEL’s thresholds that were estimated using missense variants across the whole exome^27^ and CADD’s Phred-like rank scores, which are based on the genome-wide distribution of scores for all ∼9 billion possible single nucleotide variants^18^ to establish which thresholds most accurately defined pathogenic missense variants in ALS-associated genes. When applied to a dataset restricted to missense variants classified as B/LB and P/LP in ClinVar, REVEL’s predetermined supporting pathogenicity threshold (≥0.644) captured the highest proportion of unique missense variants classified as P/LP (66.15%), corresponded most closely with the Kolmogorov-Smirnov statistic describing the maximum distance between the empirical cumulative distribution of missense B/LB and P/LP variants, and maintained high accuracy. Further, in general, for a tool to have clinical value, it should have a high NPV (∼95%) to avoid missing truly pathogenic variants and at least moderate PPV (∼50%) to minimize further clinical investigation^31^. When applied to a dataset comprising only missense B/LB and P/LP variants, the NPV for REVEL’s supporting pathogenic threshold was 97.7%, indicating that only 2.3% of truly pathogenic missense variants would receive a false-negative classification, and the PPV was 97.0%, limiting the false-positive rate to 3.0%. REVEL’s supporting pathogenic threshold also demonstrated an ability to identify benign variants with confidence, as 98.18% of unique missense variants classified as B/LB in ClinVar had a REVEL score < 0.644. Collectively, our results suggested that REVEL could be of clinical value at a cut- off of 0.644 for variants in ALS-associated genes.

In contrast, a CADD threshold ≥20.000 was highly inclusive for benign missense variants; however, it still corresponded most closely with the KS statistic and the Youden index computed from the ROC curve. A CADD threshold ≥30.000 was too restrictive, capturing only 12.31% of unique missense variants classified as P/LP in ClinVar, while a CADD threshold ≥ 20.000 captured 90.77% of unique missense variants classified as P/LP in ClinVar. Yet there were remarkable limitations regarding the relationship between CADD score and variant pathogenicity classifications for rare, missense variants in ALS-associated genes, even though the sensitivity and specificity of CADD have been shown to be high in datasets balanced for known pathogenic and benign variants^31^. Surprisingly, 64.15% of all unique missense variants had CADD scores ≥ 20.000, with 4.69% being classified as B/LB in ClinVar. It is important to note that CADD scores are distributed using a Phred-like system, such that the score of one variant is relative to the scores — and therefore deleteriousness — of all other possible variants^18^. It is well-established that rare variants are more likely to be deleterious than common variants, because of natural selection^18^, and as such we anticipate that the CADD scores of rare variants are likely to be higher than the CADD scores of common variants. Herein, we specifically selected for rare variants, which may have biased the dataset to include more variants of higher CADD score and limited the perceived accuracy of the CADD ≥ 20.000 threshold. Indeed, when CADD was applied to a dataset restricted to rare missense variants classified as B/LB and P/LP in ClinVar, the PPV at a cut-off of 20.000 was 1.01%, indicating that using this pathogenicity threshold would lead to an overwhelming overestimation of rare missense variant pathogenicity. Although CADD had an NPV of 98.3% at a cut-off of 20.000, these data ultimately suggest that the PPV of CADD is not high enough to effectively classify rare missense variants in ALS-associated genes or reduce the number of potentially pathogenic variants to those that could be efficiently followed up in clinical practice.

Concerningly, VUS carrier rates from multigene panel testing in ALS have been reported to range from 15% to 25%. These variants not only complicate genetic counseling but may prevent patient enrollment in clinical trials and are undoubtedly frustrating for patients and their families^32, 33^. Missense variants are particularly difficult to assess for pathogenicity, making up a large majority of identified VUS. Consistent with previous studies^12–15^, we observed that 20% of pALS in the ALS Knowledge Portal were carriers of at least one rare, missense VUS in an ALS-associated gene. Given the prevalence of VUS in NGS panel testing in ALS, we aimed to determine whether we could harness the *in silico* predictors and their corresponding thresholds to elucidate the pathogenicity of missense VUS in ALS-associated genes.

Enrichment analysis revealed a significantly higher frequency of VUS with a REVEL score ≥ 0.644 in pALS than controls. However, our variant-level positive yield analysis revealed that the proportion of observed REVEL-defined supporting pathogenic VUS was only marginally higher in pALS than in controls (pALS = 3.43%; controls = 2.60%). Although REVEL demonstrated strong performance distinguishing true pathogenic variants from true benign variants in our earlier analyses, the modest difference in REVEL-defined supporting pathogenic VUS identified in pALS and controls suggests the real-world value of REVEL in further evaluating VUS may be limited when applied to ALS. It is possible that the REVEL-defined supporting pathogenic VUS identified in controls may represent variants of reduced penetrance^34^, and the use of a REVEL threshold of ≥ 0.644 may remain useful in a clinical testing context for ALS; however, our analyses are limited regarding details of the true pathogenic nature of the identified VUS. Future analyses regarding the usefulness of REVEL in revealing the potential pathogenicity of VUS in ALS-associated genes would benefit from additional benchmarking using functional experimental assay evidence.

Based on the limitations of REVEL observed across all 20 ALS-associated genes, we sought to determine if the findings were specific to individual ALS-associated genes. Interestingly, pALS had a greater proportion of VUS that met REVEL’s 0.644 cut-off than controls for 15 genes, and no VUSs that exceeded REVEL’s 0.644 cut-off were observed in *SOD1*, *TARDBP*, *TUBA4A*, *UBQLN2*, and *VAPB* in controls, all of which are definitively associated with ALS^22^. Only 79 REVEL-defined supporting pathogenic VUS were observed in controls across genes that are definitively associated with ALS. The observation of supporting pathogenic missense VUS in controls may be due to false-positive classifications by REVEL or may represent cases of incomplete penetrance in the controls^34^. Furthermore, among the genes that had reached maximum genetic and experimental evidence for association with ALS by the ClinGen GCEP as of July 2023^22^ – *FUS, OPTN* [MIM: 602432]*, SOD1, SPG11, TARDBP, TBK1* [MIM: 604834]*, UBQLN2, VAPB,* and *VCP* [MIM: 601023] – all except for *SPG11* and *VCP* showed a > 2-fold increase in the proportion of observed supporting pathogenic VUS in pALS than controls (Table S1). Taken together, REVEL is most valuable for identifying VUS that are truly pathogenic in a subset of genes that are most strongly associated with ALS, which may be promising for its application in ALS clinical genetic testing as these genes are most commonly included on ALS panels^35^.

Unsurprisingly, the performance of CADD in evaluating the pathogenicity of VUS in ALS-associated genes trailed behind REVEL. Enrichment analysis revealed no significant difference in the frequency of VUS with a CADD score ≥ 20.000 in pALS compared to controls.

Additionally, the proportion of observed CADD-defined supporting pathogenic missense VUS was nearly equal between pALS and controls (pALS = 23.36%; controls = 23.88%). At a gene-specific level, a remarkably high proportion of VUSs exceeded CADD’s threshold in both pALS and controls across all 20 ALS-associated genes. Based on our findings, CADD is of limited value for refining pathogenicity prediction of missense VUSs in ALS-associated genes given the exceedingly high false-positive rate for the control cohort.

We conclude that a REVEL score ≥0.644 for rare, missense variants in ALS-associated genes is likely sufficient to identify variants of further pathogenic interest in a clinical and research setting, especially for genes which are most strongly associated with ALS, including *FUS, OPTN, SOD1, TARDBP, TBK1, UBQLN2,* and *VAPB*. In contrast, employing CADD’s ≥ 20.000 threshold demonstrated a remarkably low positive predictive value for identifying truly pathogenic missense variants in ALS-associated genes; thus, the use of CADD may lead to a gross overestimation of supporting pathogenic variants in ALS. Our findings highlight the need for improved tools that are specific to disease-associated genes and molecular pathways, to accurately predict the pathogenicity of VUSs in multifactorial, late-onset disorders such as ALS.

## Supplemental Information

Supplemental Information includes 8 figures and 4 tables.

## Data Availability

ALS Knowledge Portal data is available through the Neurodegenerative Disease Knowledge Portal (https://ndkp.hugeamp.org). Project MinE sequencing consortium data is available through the Project MinE Databrowser (http://databrowser.projectmine.com).

http://databrowser.projectmine.com

## Acknowledgments

MRF is supported by the Canadian Institute of Health Research Canada Graduate Scholarships-Master’s program and the Fonds de Recherche Santé Québec Master’s training program. AAD is supported by the Canadian Institute of Health Research Banting Postdoctoral Fellowship program. SMKF is supported by grants from ALS Canada, Brain Canada, The Michael J. Fox Foundation, and The Montreal Neurological Institute-Hospital.

## Author contributions

M.R.F.: conceptualization, data curation, methodology, formal analyses, visualization, writing of the original draft, and review/editing of the manuscript; A.A.D: conceptualization, data curation, methodology, and review/editing of the manuscript; S.M.K.F.: supervision, conceptualization, methodology, and review/editing of the manuscript.

## Web Resources

ALS ClinGen Gene Curation Expert Panel, https://search.clinicalgenome.org/kb/affiliate/10096?page=1&size=25&search=

ClinVar, https://www.ncbi.nlm.nih.gov/clinvar/

Combine Annotation Dependent Depletion (CADD), https://cadd.gs.washington.edu

Emsebl Variant Effect Predictor (VEP), https://useast.ensembl.org/info/docs/tools/vep/index.html

Neurodegenerative Disease Knowledge Portal (formerly known as the ALS Knowledge Portal), https://ndkp.hugeamp.org

OMIM, http://www.omim.org/

Project MinE Data Browser, http://databrowser.projectmine.com

R, https://www.r-project.org

Rare Exome Variant Ensemble Learner (REVEL), https://sites.google.com/site/revelgenomics/

ROCit R package, https://cran.r-project.org/web/packages/ROCit/vignettes/my-vignette.html

