## Supplemental Information for "Evaluating the utility of REVEL and CADD for interpreting variants in amyotrophic lateral sclerosis genes"

### Supplemental Figures

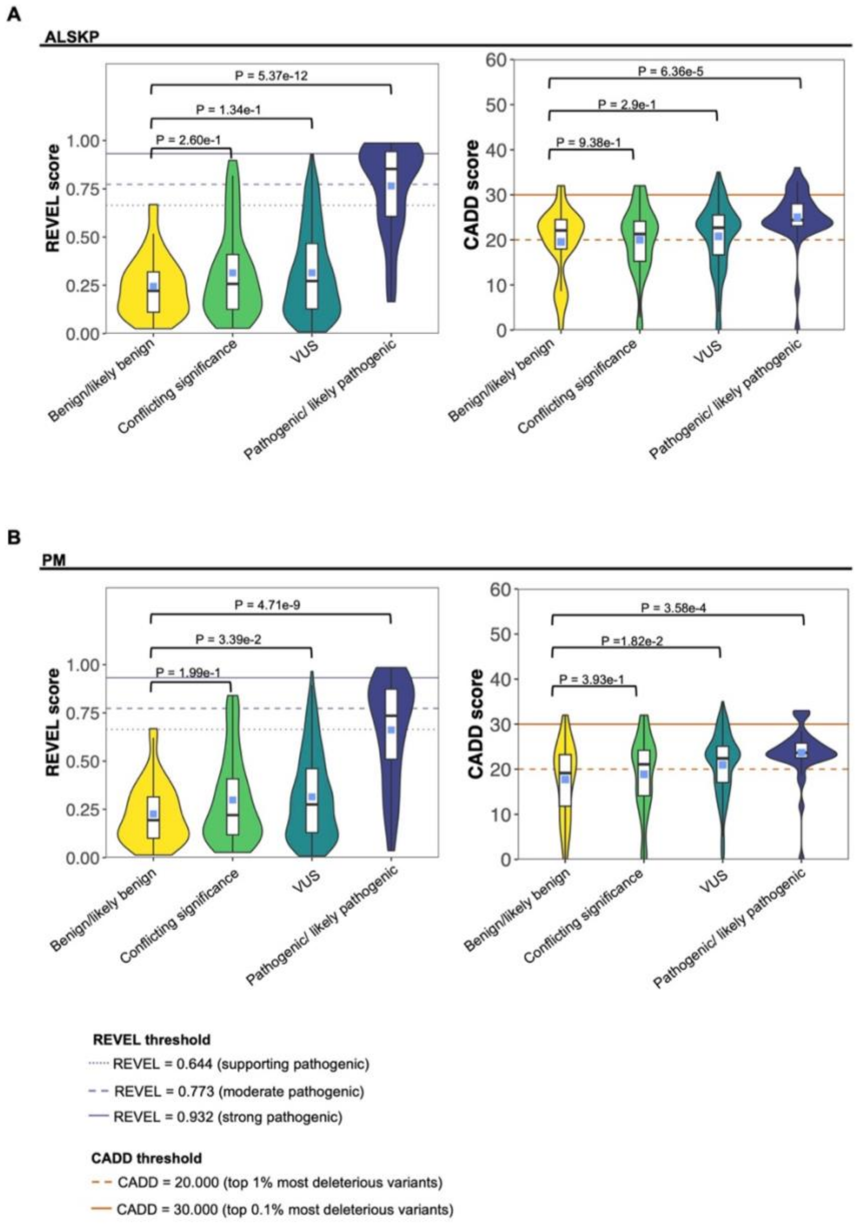


**Figure S1. Distribution of REVEL and CADD scores for rare, missense variants in ALS-associated genes categorized by ClinVar classification.** REVEL and CADD scores for unique variants identified in ClinGen definitive or moderate ALS-associated genes in the **A)** ALS Knowledge Portal (ALSKP) dataset and **B)** Project Mine ALS Sequencing Consortium (PM) dataset, stratified by ClinVar classification. Wilcoxon Rank-sum tests were conducted to compare the mean REVEL or CADD scores of each ClinVar category to the mean REVEL or CADD score of benign/likely benign variants. Abbreviations: VUS, variants of uncertain significance.

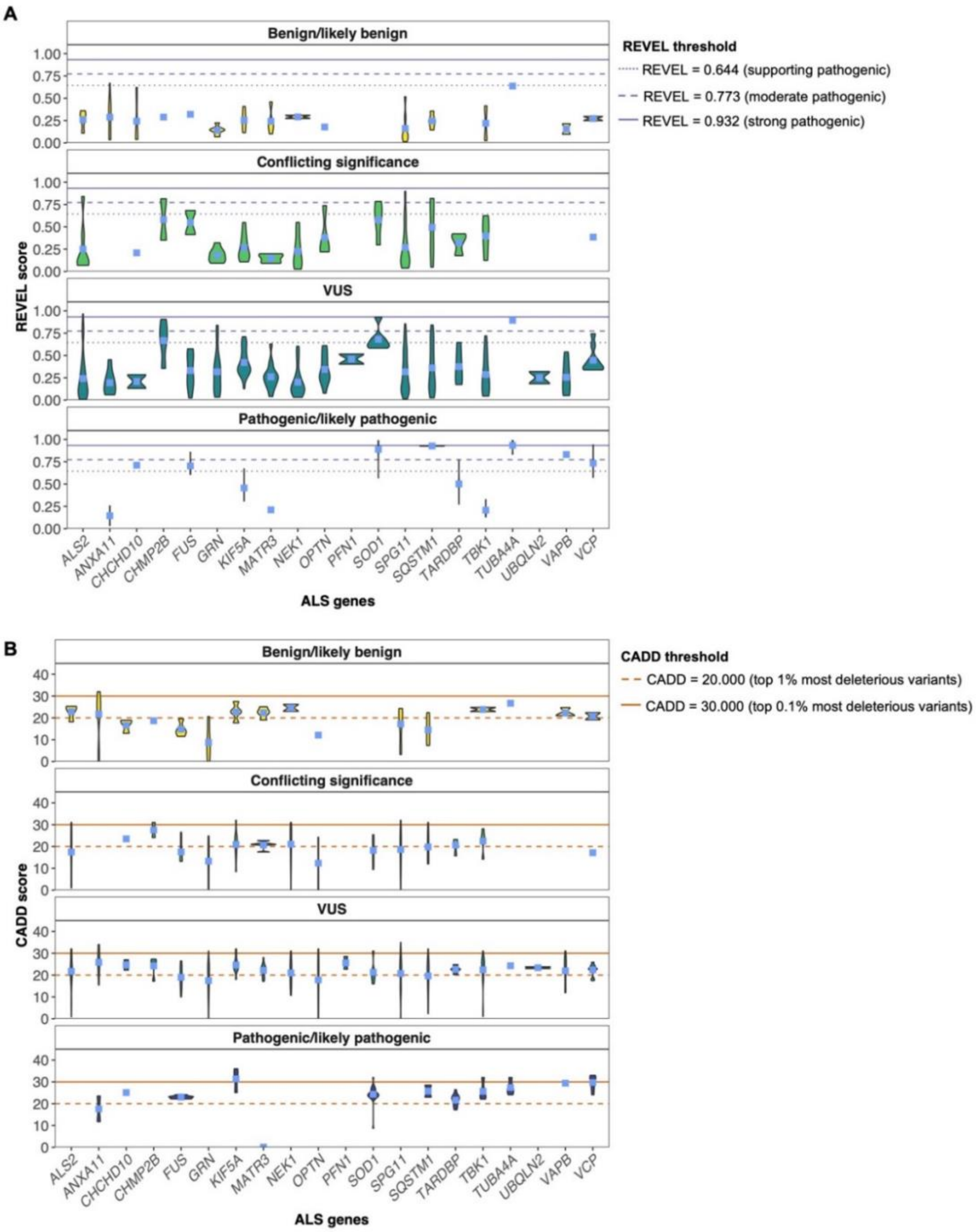


**Figure S2. CADD and REVEL scores for rare, missense variants in ALS-associated genes, stratified by gene and ClinVar classification. A)** Distribution of REVEL scores for all unique missense variants observed in ClinGen definitive or moderate ALS-associated genes across the ALS Knowledge Portal (ALSKP) and Project Mine ALS Sequencing Consoritum (PM) datasets. **B)** Distribution of CADD scores for all unique missense variants observed in ClinGen definitive or moderate ALS-associated genes across the ALSKP and PM datasets.

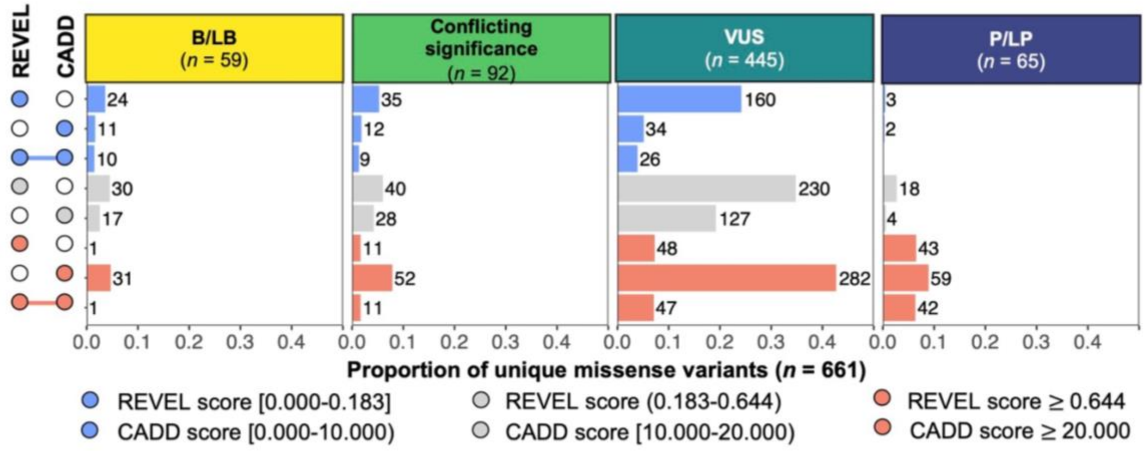


**Figure S3. Number of missense variants in ClinGen definitive or moderate ALS-associated genes observed in each ClinVar category, stratified by REVEL or CADD score.** The x-axis shows the proportion of all unique missense variants observed. The number of unique variants in the respective category is listed beside each bar. The upset plot on the y-axis indicates which REVEL or CADD bins are captured by the respective bar. Abbreviations: B/LB, benign/likely benign; P/LP, pathogenic/likely pathogenic; VUS, variant of uncertain significance.

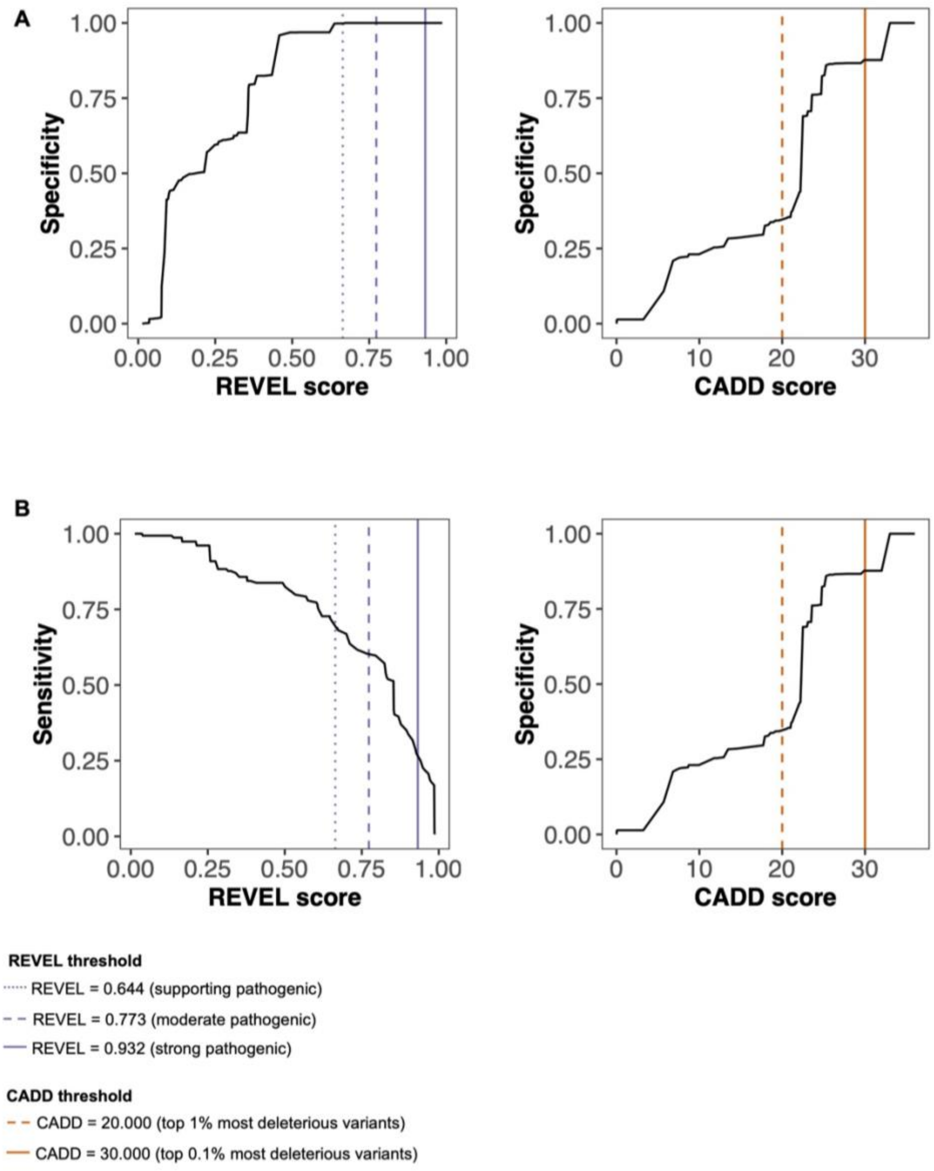


**Figure S4. Sensitivity and specificity of REVEL and CADD scores for rare, missense variants in ClinGen definitive or moderate ALS-associated genes. A)** Specificity of REVEL and CADD for identifying variants classified as benign/likely benign in ClinVar in a dataset restricted to missense variants classified as benign/likely benign and pathogenic/likely pathogenic in ClinVar. **B)** Sensitivity of REVEL and CADD for identifying variants classified as pathogenic/likely pathogenic in ClinVar in a dataset restricted to missense variants classified as benign/likely benign and pathogenic/likely pathogenic in ClinVar.

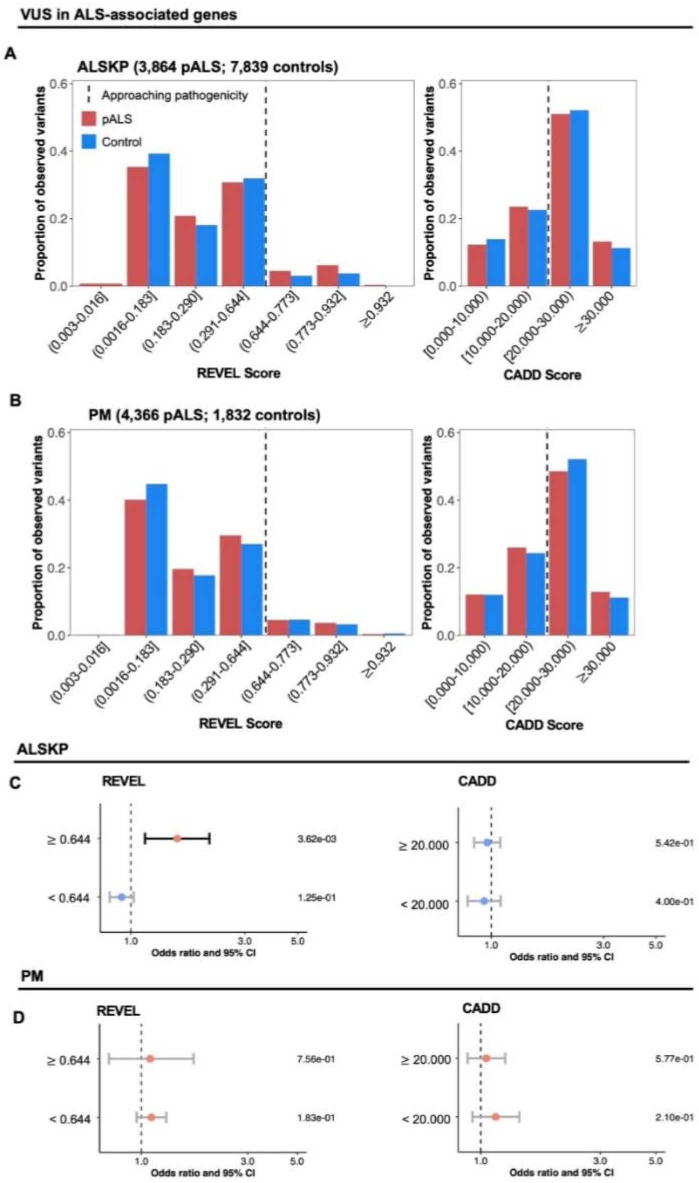


**Figure S5. Distribution of rare, missense VUS in ClinGen definitive or moderate ALS-associated genes seen in pALS and controls.** Variants categorized as variants of uncertain significance (VUS) in ClinVar, conflicting in ClinVar, or absent from ClinVar are collectively referred to as VUS. **A)** Proportion of observed missense VUS seen in the ALS Knowledge Portal (ALSKP) dataset, stratified by ALS status and binned REVEL or CADD score. **B)** Proportion of observed missense VUS seen in the Project MinE ALS Sequencing Consortium (PM) dataset, stratified by ALS status and binned REVEL or CADD score. **C)** Fisher's Exact tests investigating whether people with ALS (pALS) from the ALSKP dataset were enriched for missense VUS that had REVEL or CADD scores that exceeded their respective pathogenicity thresholds (REVEL ≥ 0.644; CADD ≥ 20.000) compared to controls. **D)** Fisher's Exact tests investigating whether pALS from the PM dataset were enriched for missense VUS that had REVEL or CADD scores that exceeded their respective pathogenicity thresholds (REVEL ≥ 0.644; CADD ≥ 20.000) compared to controls. P-values corresponding to each variant category are shown on the right of the plot. Abbreviations: CI, confidence interval.

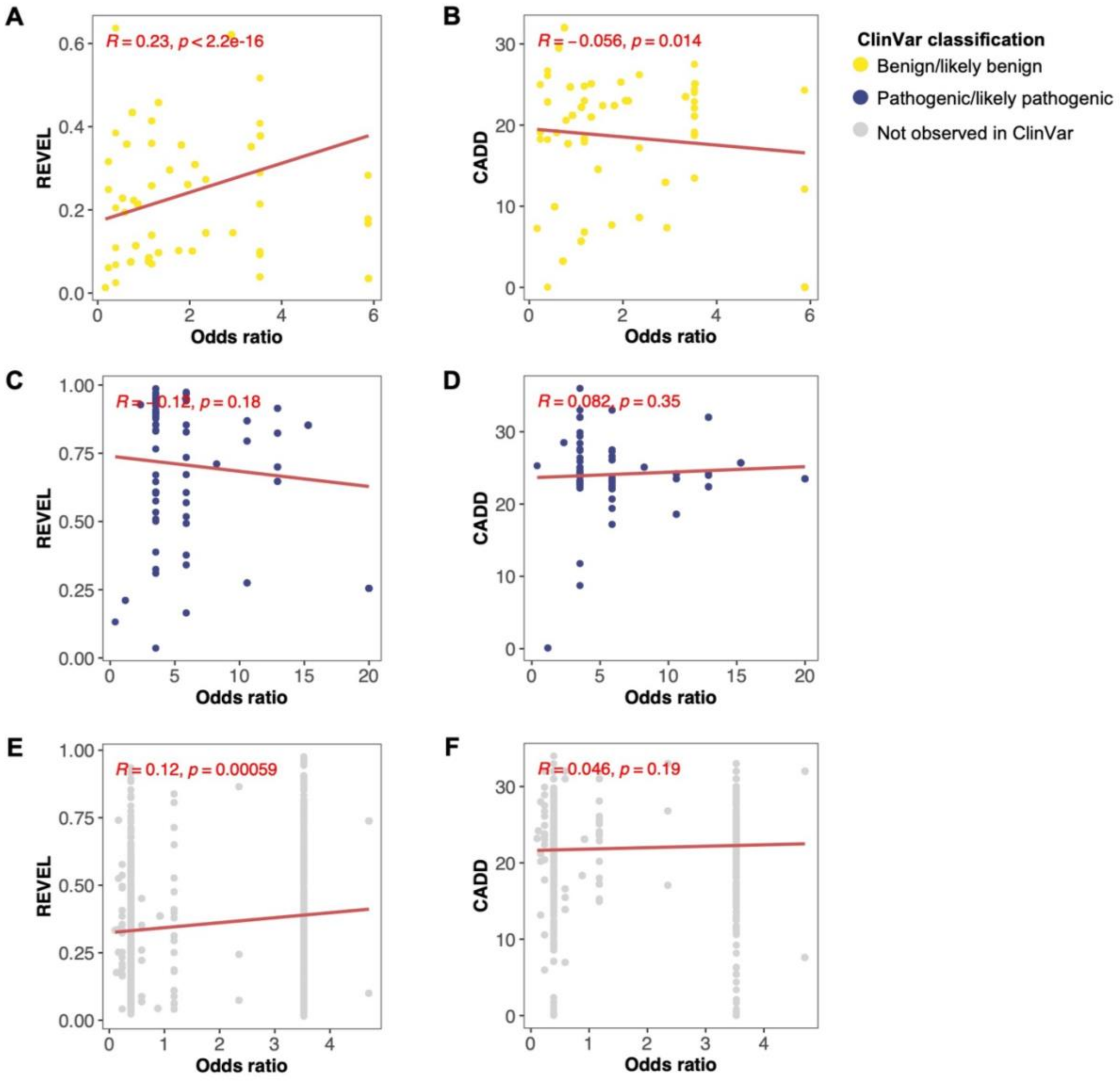


**Figure S6. REVEL and CADD scores for rare, missense variants in ClinGen definitive or moderate ALS-associated genes according to variant odds ratio. A)** Correlation between REVEL score and variant odds ratio for missense variants classified as benign/likely benign (B/LB) in ClinVar. **B)** Correlation between CADD score and variant odds ratio for missense variants classified as B/LB in ClinVar. **C)** Correlation between REVEL score and variant odds ratio for missense variants classified as pathogenic/likely pathogenic (P/LP) in ClinVar. **D)** Correlation between CADD score and variant odds ratio for missense variants classified as P/LP in ClinVar. **E)** Correlation between REVEL score and variant odds ratio for missense variants that are absent from ClinVar. **F)** Correlation between CADD score and variant odds ratio for missense variants that are absent from ClinVar. Only variants that had an odds ratio within three standard deviations of the mean are shown and were included when computing Pearson's correlation.

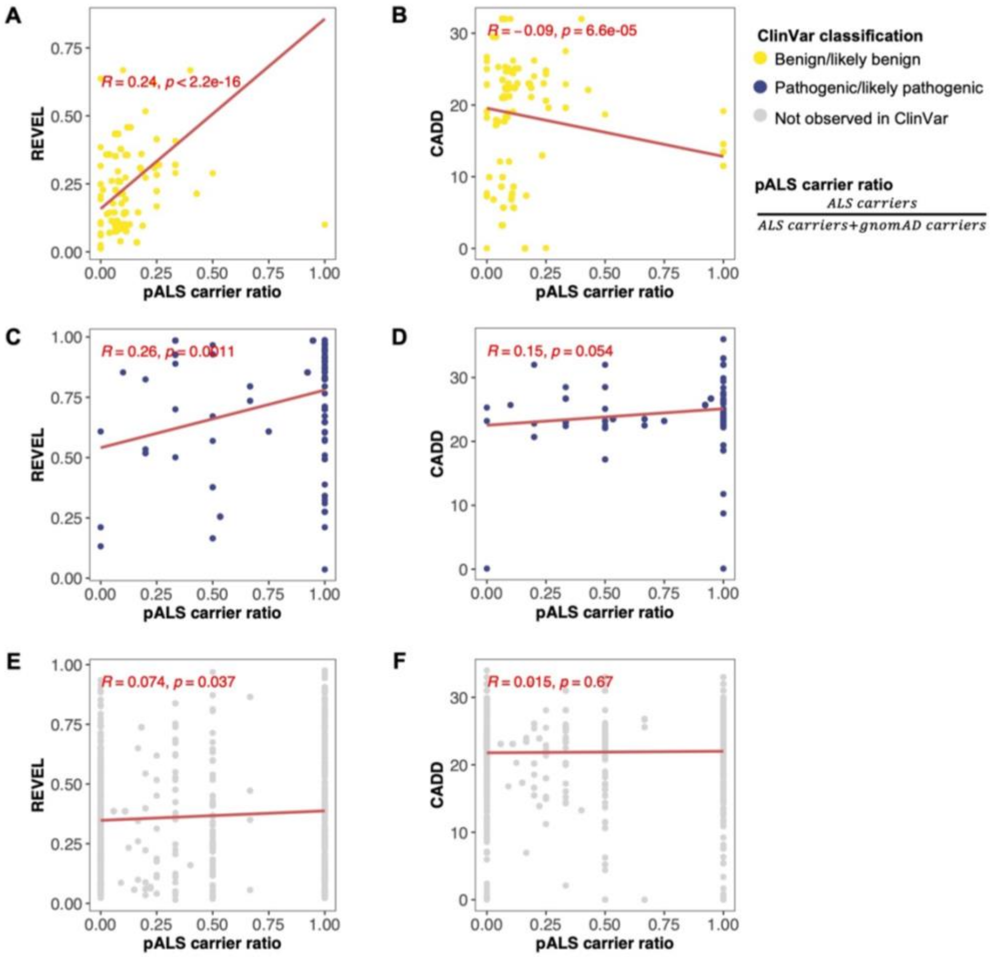


**Figure S7. REVEL and CADD scores for rare, missense variants in ClinGen definitive or moderate ALS-associated genes according to pALS carrier ratio. A)** Correlation between REVEL score and variant carrier ratio in pALS (pALS carrier ratio) for missense variants classified as benign/likely benign (B/LB) in ClinVar. **B)** Correlation between CADD score and pALS carrier ratio for missense variants classified as B/LB in ClinVar. **C)** Correlation between REVEL score and pALS carrier ratio for missense variants classified as pathogenic/likely pathogenic (P/LP) in ClinVar. **D)** Correlation between CADD score and pALS carrier ratio for missense variants classified as P/LP in ClinVar. **E)** Correlation between REVEL score and pALS carrier ratio for missense variants that are absent from ClinVar. **F)** Correlation between CADD score and pALS carrier ratio for missense variants that are absent from ClinVar.

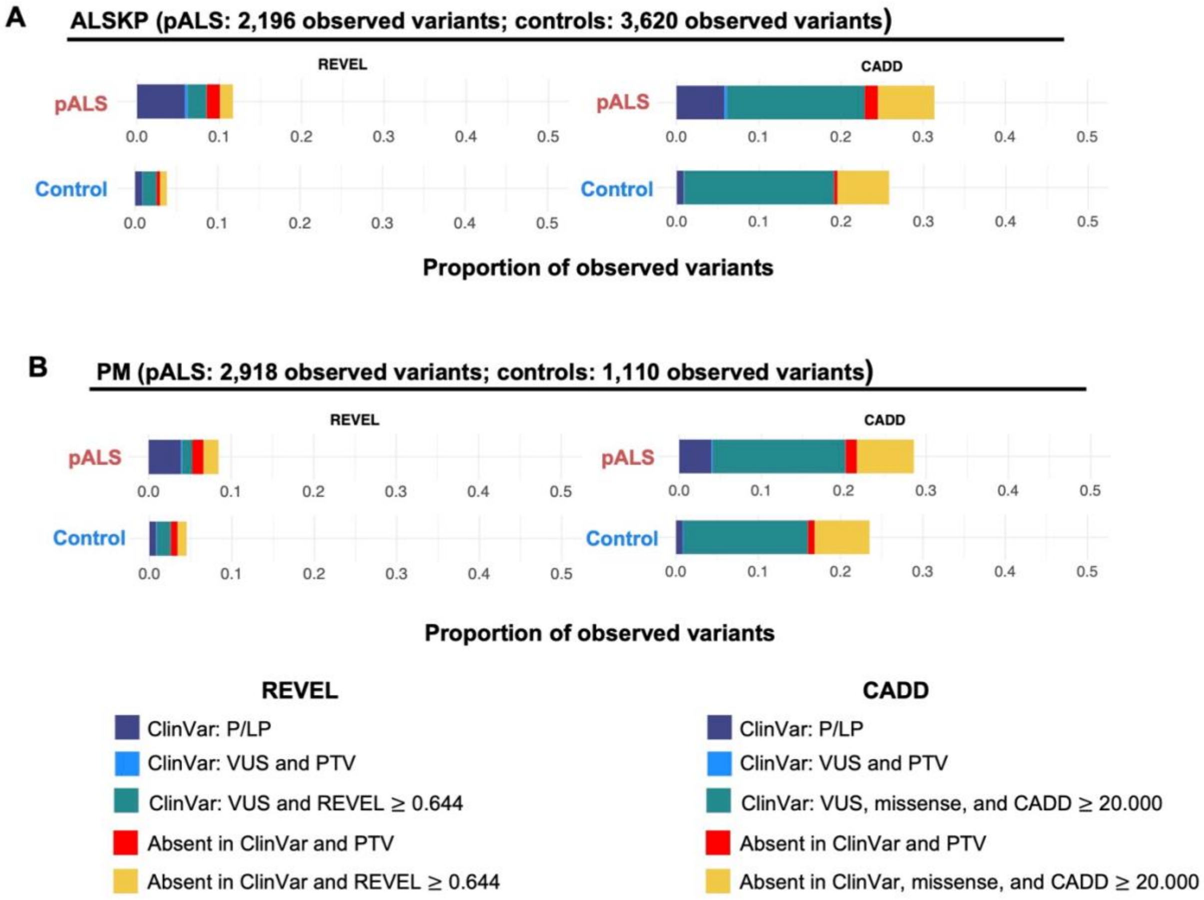


**Figure S8. Proportion of observed variants identified as being supporting pathogenic in pALS and controls.** Rare, coding variants in ClinGen definitive or moderate ALS-associated genes were considered supporting pathogenic if they were either pathogenic/likely pathogenic (P/LP) in ClinVar, protein-truncating variant (PTV) of uncertain significance in ClinVar, missense VUS in ClinVar with REVEL $\geq$ 0.644/CADD $\geq$ 20.000, PTV absent from ClinVar, or a missense variant absent from ClinVar with REVEL $\geq$ 0.644/CADD $\geq$ 20.000. **A)** Proportion of observed variants in people with ALS (pALS) and controls from the ALS Knowledge Portal (ALSKP) dataset that were identified as being supporting pathogenic by ClinVar and REVEL (left) or ClinVar and CADD (right). **B)** Proportion of observed variants in pALS and controls from the Project MinE ALS Sequencing Consortium (PM) dataset that were identified as being supporting pathogenic by ClinVar and REVEL (left) or ClinVar and CADD (right).

### Supplemental Tables

**Table S1. Gene-disease validity classifications from the ALS ClinGen Gene Curation Expert Panel for ALS-associated genes.**

| Gene | ALS ClinGen GCEP classification | Genetic evidence | Experimental evidence | Last evaluation |
|---|---|---|---|---|
| *ALS2* | Definitive | 12.00 | 3.50 | 02/14/2023 |
| *ANXA11* | Definitive | 12.00 | 5.50 | 11/12/2021 |
| *CHMP2B* | Definitive | 6.40 | 6.00 | 07/12/2022 |
| *FUS* | Definitive | 12.00 | 6.00 | 10/12/2021 |
| *GRN* | Definitive | 12.00 | 3.00 | 05/25/2023 |
| *KIF5A* | Definitive | 12.00 | 2.00 | 05/26/2022 |
| *NEK1* | Definitive | 12.00 | 1.50 | 04/29/2022 |
| *OPTN* | Definitive | 12.00 | 6.00 | 05/10/2022 |
| *PFN1* | Definitive | 10.70 | 6.00 | 04/22/2021 |
| *SOD1* | Definitive | 12.00 | 6.00 | 07/13/2021 |
| *SPG11* | Definitive | 12.00 | 6.00 | 06/07/2023 |
| *TARDBP* | Definitive | 12.00 | 6.00 | 07/27/2021 |
| *TBK1* | Definitive | 12.00 | 6.00 | 11/03/2022 |
| *UBQLN2* | Definitive | 12.00 | 6.00 | 04/13/2021 |
| *VAPB* | Definitive | 12.00 | 6.00 | 12/15/2021 |
| *VCP* | Definitive | 12.00 | 6.00 | 12/23/2021 |
| *CHCHD10* | Moderate | 3.20 | 4.50 | 09/13/2022 |
| *MATR3* | Moderate | 3.50 | 4.50 | 09/23/2021 |
| *SQSTM1* | Moderate | 5.60 | 6.00 | 12/13/2022 |
| *TUBA4A* | Moderate | 4.10 | 2.00 | 09/14/2021 |

All ALS-associated genes with definitive or moderate gene-disease validity classifications as of July 2023 are shown and were included in the analysis. The maximum number of genetic evidence and experimental evidence points are 12.00 and 6.00, respectively. Abbreviations: GCEP, Gene curation expert panel.

**Table S2. Recategorization of ClinVar classifications.**

| Recategorized ClinVar classification | Original ClinVar Classification |
|---|---|
| Other | Other, Affects, confers sensitivity |
| Conflicting significance | Conflicting interpretations of pathogenicity, Conflicting interpretations of pathogenicity; association, Conflicting interpretations of pathogenicity; other, Conflicting interpretations of pathogenicity; risk factor, Conflicting interpretations of pathogenicity; drug response, Conflicting interpretations of pathogenicity; association; risk factor |
| Variant of uncertain significance | Uncertain significance, Uncertain significance; risk factor, Uncertain significance; association, Uncertain significance; other |
| Protective | Protective |
| Benign/likely benign | Likely benign, Benign, Benign/Likely benign, Benign/Likely benign; other, Benign/Likely benign; drug response, Benign; other, Benign; confers sensitivity, Benign/Likely benign; risk factor, Benign; risk factor, Likely benign; other |
| Drug response | Drug response, drug response; risk factor |
| Association | Association, association; risk factor |
| Risk factor/likely risk factor | Risk factor, Likely risk allele, Affects; risk factor |
| Pathogenic/likely pathogenic | Pathogenic, Likely pathogenic, Pathogenic/Likely pathogenic, Likely pathogenic; risk factor, Pathogenic/Likely pathogenic; other, Pathogenic; risk factor, Pathogenic/Likely pathogenic; risk factor, Pathogenic; other, Pathogenic; drug response, Likely pathogenic; drug response, Likely pathogenic; other, Pathogenic; Affects |
| Not observed in ClinVar | *NA*, not provided, no interpretation for the single variant |

**Table S3. Transcript identifiers for ALS-associated genes used as input into Variant Effect Predictor.**
*Table_S3.xlsx*

**Table S4. Rare, coding variants in ALS-associated genes identified as supporting pathogenic.** Variants were considered supporting pathogenic if they were either a pathogenic/likely pathogenic (P/LP) in ClinVar, protein-truncating variant (PTV) of uncertain significance in ClinVar, missense variant of uncertain significance (VUS) in ClinVar with REVEL $\geq$ 0.644, PTV absent from ClinVar, or a missense variant absent from ClinVar with REVEL $\geq$ 0.644.
*Table_S4.xlsx*
